# Assessing the impact of absence of coordination in malaria intervention strategies: a modelling study

**DOI:** 10.64898/2026.06.03.26354857

**Authors:** Younes Iggidr, Nick Ruktanonchai, Bilal Benhana, Valérian Turbé, Billy Bauzile, Abigail Ward, Justin Cohen, Emilie Pothin, Clara Champagne

## Abstract

Malaria control programs are increasingly tailored at subnational scales; however, neighboring areas remain connected through human mobility, allowing parasite importation that may undermine independently timed interventions. Although the spatial targeting of control has been the focus of extensive research, the epidemiological consequences of temporal misalignment in intervention deployment across interconnected regions remain to be elucidated. We investigate how asynchronous timing of malaria interventions affects transmission dynamics using a two-patch susceptible–infected–susceptible metapopulation model. We compare synchronous and asynchronous intervention schedules and quantify their impact using measures of excess cumulative incidence attributable to asynchrony. The measure that will be used for this purpose is referred to as Asynchrony Induced Growth (AIG).

Across a range of 10,000 parameter combinations, asynchronous implementation has been observed to result in a heightened incidence compared to synchronized deployment, though the impact is typically negligible in most endemic settings. Sensitivity analyses indicate that the impact is most significant when interventions are highly effective, infectious duration is brief, and transmission intensity approaches the elimination threshold. In such circumstances, asynchrony has the potential to substantially inflate case numbers, delay transmission interruption, or even prevent elimination entirely. In illustrative scenarios that reflect realistic settings, synchronizing interventions has been shown to avert large numbers of infections and shorten elimination timelines by years to decades.

These findings demonstrate that, beyond spatial targeting, temporal coordination of interventions across connected areas can meaningfully enhance malaria control and elimination. Coordinated timing may be particularly valuable for cross-border or near-elimination programs and should be considered in operational planning and resource allocation.

## Introduction

Like many infectious diseases, malaria transmission is influenced by a large variety of factors, linked to the environment, human behavior and the implementation of control interventions (Cohen et al. 2017; Villena et al. 2024). The combination of these factors makes each geographical setting a unique epidemiological context. Thus, there is an increasing emphasis on taking these geographical specificities into account to design intervention strategies tailored to the local context at the national and subnational levels (World Health Organization 2018; World Health Organization 2025; Thawer et al. 2020; Diallo et al. 2024). However, these initiatives often consider countries, provinces, or districts as independent entities.

In reality, geographical areas are not isolated from one another. Pathogens know no borders and travel across space through the movement of people and vectors. Human geographical mobility, in particular, can substantially impact malaria transmission dynamics. Mobility can hinder elimination efforts by sustaining transmission in areas where local conditions alone would suffice to eliminate the disease (Ruktanonchai et al. 2016; Wesolowski et al. 2012; Ihantamalala et al. 2018; Champagne et al. 2022; Das et al. 2023). Cross-border malaria has been proven to be a major obstacle for malaria elimination (Wangdi et al. 2015). Additionally, the impact of intervention strategies can differ in the presence of connectivity compared to what would be achieved if areas were isolated (Das et al. 2022; Champagne et al. 2024; Silal et al. 2015; Tun et al. 2021; Nikolov et al. 2016; Agusto et al. 2021; Saucedo and Tien 2022). Therefore, the optimal intervention strategy in a given place may depend on the interventions deployed in neighboring areas (Khadka et al., 2018). Indeed, collaborative bi-national and multinational initiatives have led to significant reduction in malaria burden and mortality, in particular along border regions (Fambirai et al. 2022). Conversely, malaria elimination efforts have been negatively impacted by connectivity and importation in places such as the DRC (Carrel et al., 2015) and Peru (Carrasco-Escobar et al., 2024), and this issue has been highlighted as a critical issue for elimination campaigns generally (Sturrock et al., 2015).

Typically, accounting for connectivity has meant spatial optimization of interventions, such as making sure to target “source” areas (Moonen et al., 2010) or hotspots (Bousema et al., 2012) to prevent importation into downstream locations. Recent research has shown that temporal optimization timing can also have important consequences for disease control. For example, during the Covid-19 pandemic, (Ruktanonchai et al. 2020) showed that if different countries in the European Union came out of lockdown at different times, there would be many more infections and hospitalizations from Covid-19 than if countries and locations had come out of lockdown in a coordinated manner.

Metapopulation models have been commonly used to address how spatial and temporal variation affect disease persistence. By representing regions as sets of communities or “patches” that are interconnected, these models account for disease spread across patches (Cosner et al. 2009; Prosper et al. 2012). Recently, (Kortessis et al. 2024) used a metapopulation model to extensively describe a theoretical effect called “inflationary effect”, whereby spatiotemporal heterogeneity between areas – e.g. driven by spatially asynchronous interventions – paired with host movement between areas can inflate the resulting number of infected hosts, compared to a scenario with higher synchrony. Similarly, (Katriel 2022) describes a similar phenomenon in a time-periodic environment, called “DIG” (Dispersal Induced Growth), where all patches are sinks, but the overall disease growth rate is positive and states the “all-sink case” theorem, giving mathematical conditions on patches growth rates for this to occur.

Diseases like malaria, which exhibit strong spatiotemporal heterogeneity in transmission rates, are a useful setting for this research. Numerous studies have examined the effects of spatial targeting of intervention effort (Walker et al., 2016, Runge et al., 2022, Ozodiegwu et al., 2023), including field studies investigating effectiveness (Bousema et al., 2016). To our knowledge, the effects of intervention timing on malaria have not yet been specifically investigated. It is unclear how spatiotemporal changes in intervention effort could impact malaria control, because it is a disease with different characteristics than respiratory viruses like SARS-CoV-2, exhibiting high endemicity levels, long duration of infectiousness, and different interventions deployed for prevention and treatment. These characteristics make understanding the importance of timing critical, particularly because interventions are already deployed at a variety of scales and cycles, based on the projected protective duration of the tools used (e.g. 3-year mass campaigns for Insecticide-treated nets, 6 months cycles for Indoor residual spraying) or based on seasonal patterns (e.g. seasonal malaria chemoprevention). These overlapping factors create a high potential for asynchrony between interventions and, consequently, for spatial and temporal targeting of intervention effort.

The aim of this study is to assess how a lack of coordination in the timing of malaria control interventions can influence disease dynamics. Using a two-patch metapopulation model, we quantify the epidemiological impact of intervention asynchrony through newly defined metrics. We then explore how this impact varies across a broad range of malaria transmission settings, using sensitivity analyses to identify the most influential factors. Finally, we illustrate the relevance of our findings through case-study scenarios inspired by real-world contexts. Together, these analyses aim to determine whether the timing of interventions across connected areas should be intentionally coordinated to enhance malaria control and elimination efforts.

## Methods

### Two-patch metapopulation model of malaria dynamics with control

The chosen model for this study is derived from (Ruktanonchai et al. 2016). It is an Susceptible-Infected-Susceptible (SIS) model with two patches, with *λ*_*i*_ = *rR*_0,*i*_ being the force of infection *i, R*_0,*i*_ the basic reproduction number in patch *i* and *r* the recovery rate. Each patch *i* has a constant population size *N*_*i*_ . We model connectivity using the Lagrangian approach described in (Cosner et al. 2009), i.e. we consider that each day for all *i, j* in {1, …*n*} individuals from patch *i* spend a proportion of time *p*_*ij*_ visiting patch *j*. Thus, individuals can be infected either locally or while visiting another patch, and they can infect the population of another patch while visiting. Moreover, we model an intervention in each patch *i* by a function ω_*i*_ ∈ [0, 1], with 1 representing perfect control eliminating any transmission in the patch and 0 representing the total absence of control. We do not explicitly consider vectors in this model: assuming that mosquitoes have a shorter generation time than humans, one can consider vector dynamics to be at equilibrium and approximate parasite dynamics at low endemicity with a human to-human transmission model, where the transmission parameter is proportional to the vectorial capacity (see equation 21 in Smith and McKenzie (2004)). The model is represented by the following differential expression where *X*_*i*_ is the prevalence rate in patch *i* (all notations are provided in Table 1):

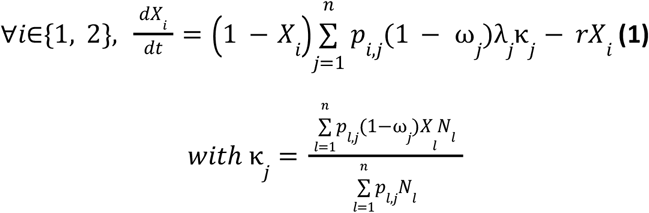

**Table 1:** Range of the parameters used for simulations.

| Notation | Description | Unit | Range used in model |
| --- | --- | --- | --- |
| $r$ | Recovery rate | $days^{-1}$ | $\left(\frac{1}{60}, \frac{1}{200}\right)$ |
| $R_{0,i}$ | Basic reproduction number in area $i$ | - | (0.9, 2.2) |
| $\omega_i$ | Effectiveness of the intervention in area $i$ | - | (0, 0.5) |
| $p_{ij}$ | Proportion of time people from area $i$ spend visiting area $j$ | - | (0, 0.5) |
| $t_I$ | Duration of one intervention | years | {0.5, 1, 1.5, ..., 5} |

**Table 2:** Number of cases and time to transmission interruption for both scenarios, in total and in each area.

|  | Cumulated annual incidence per 1000 people before transmission interruption | Time to transmission interruption (in years) | Areas | Cumulated annual incidence per 1000 people before transmission interruption in each area | Time to transmission interruption in each area (in years) |
| --- | --- | --- | --- | --- | --- |
| Synchronous scenario | 914.7 | 46 | Area 1 | 457.35 | 46 |
|  |  |  | Area 2 | 457.35 | 46 |
| Asynchronous scenario | 1862.24 | 78 | Area 1 | 741.72 | 74 |
|  |  |  | Area 2 | 1120.51 | 78 |

The incidence at day t is defined as 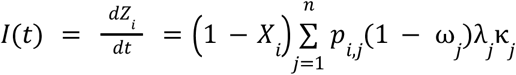 with *Z*_*i*_ being the cumulative incidence at day t.

We initialise our model at endemic equilibrium in the absence of intervention (ω_1_ = ω_2_ = 0) with prevalence 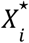 (shown in Appendix A to be unique, positive and is globally asymptotically stable). We compute 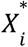 in terms of the other parameters by solving numerically System A shown in Appendix A, using Newton method implemented in the nleqslv package (Hasselman 2023).

To explore the effect of asynchrony, we then deploy interventions in the model under two scenarios. Let *t*_*I*_ be the duration of the intervention. A cycle of interventions is organized as follows : *t*_*I*_ years of control then a break of *t*_*I*_ years, i.e. we choose a value ω _*i*_ representing the strength of the intervention and therefore for all *t* in [0, 365*t*_*I*_ ], ω_*i*_ (*t*) = ω_*i*_ while for all *t* in [365*t*_*I*_, 730*t*_*I*_ ], ω_*i*_ (*t*) = 0. In the “synchronous scenario”, the interventions have identical deployment schedules in both patches. In the “asynchronous scenario”, the timing of interventions differs between the two patches: when one patch implements an intervention, the other one stops. These scenarios are illustrated in Figure 1. Notably, patch 1 always follows the same intervention schedule in both scenarios—any difference observed arises solely from the timing in patch 2.

**Figure 1:**
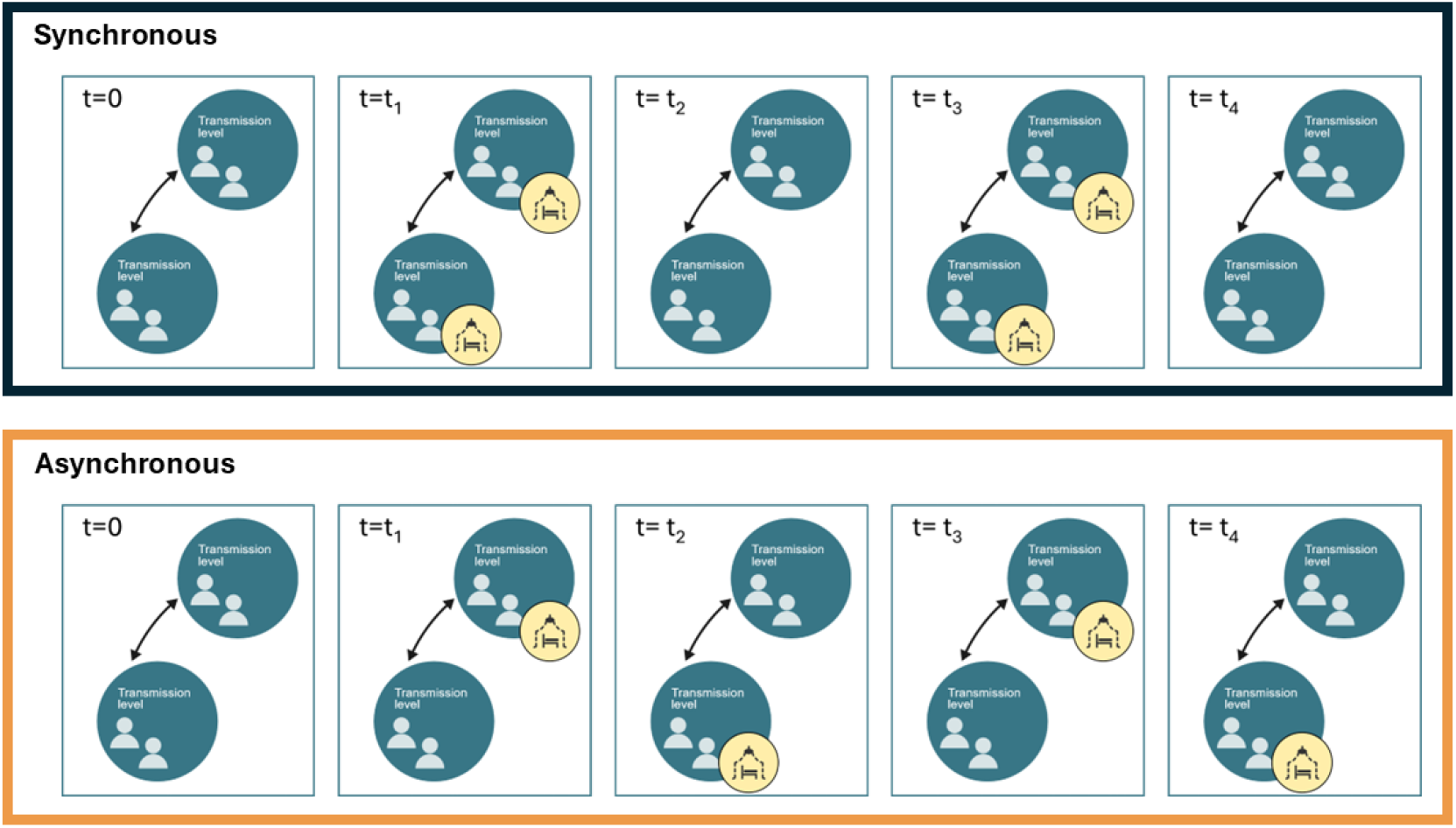
Schematic representation of the scenarios considered. Area 1 is represented by the dark blue upper bubble while Area 2 is represented by the dark blue lower bubble. In the synchronous scenario (top), interventions, represented by yellow bubbles, occur simultaneously in both areas ; in the asynchronous scenario (bottom), interventions alternate between areas and are out of phase. The two-headed arrow represents human movement between the two patches.

The model is implemented in R using the odin package (FitzJohn et al. 2025).

### Metrics

To measure the impact of asynchronous interventions, we define the following metrics.

**Definition (Asynchrony Induced Growth, AIG)**: Let *I*_*sync*_ (*t*) and *I*_*async*_ (*t*) denote the incidence at day t in the synchronous and asynchronous scenarios respectively. The *AIG* on *n* days is defined as 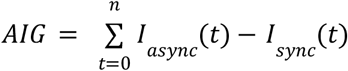.

This gives a metric in terms of additional incidence burden in the asynchronous case. As AIG is susceptible to scaling effects regarding the intensity of the synchronous epidemic, we also defined the Asynchrony Induced Growth Rate.

#### Definition (Asynchrony Induced Growth Rate, AIGR)

With the same notations, and assuming that there exists a *t*_0_ ∈{0, …, *n*} such as *I*_*sync*_ (*t*_0_ ) > 0, AIGR is defined as the following rate:

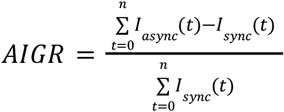

While this indicator can be computed for both patches, we will only consider its value for the first patch in most analyses. As previously mentioned, the first patch receives the exact same intervention schedule in both scenarios, so any change in the metric would only be due to changes in the timing of interventions in the second patch. This is done without loss of generality, as the two patches could be swapped.

We will look at detecting transmission interruption (or lack of interruption because of asynchrony). We then define transmission interruption in this article as having an annual incidence of less than 1 case per 100,000 persons per year.

### Uncertainty and sensitivity analysis

We investigated the impact of intervention asynchrony with our model by computing the AIG metric (AIGR giving the same results, we chose AIG to express the results in terms of added cases per year per 1000 people) over a wide range of scenarios, to identify both the magnitude of the effect and the most influential parameters. For this, we sampled 10,000 sets of parameters with a Latin Hypercube using the minimax criterion (Lin and Tang 2014), over ranges indicated in Table 1. We chose a recovery time between 60 days and 200 days to represent diverse malaria parasites (60 days for *P. vivax* according to (McKenzie et al. 2002) and 200 days for *P. falciparum* according to (Felger et al. 2012)). An upper bound of 2.2 for the reproduction number is inspired by (Ruktanonchai et al. 2016), to be larger than for example the *R*_0_ in Namibia estimated in the cited article. We simulated our model with each parameter set over a period comprising 3 intervention cycles, a cycle being 2*t*_*I*_ years (*t*_*I*_ years when the intervention is deployed, followed by *t*_*I*_ years where the intervention is stopped). The AIG is computed only on 6*t*_*I*_ years, and the AIG per year is the AIG divided by 6*t*_*I*_.

The relative contribution of the parameters to the variability of the Asynchrony Induced Growth is explored by variance decomposition, calculating Sobol indices with the Sobol function of the sensitivity R package (Iooss et al. 2024).

### Decision tree method

We investigated the characteristics of the scenarios leading to the highest impact of asynchrony, namely the scenarios with the 10% highest AIG values. To characterize them, we constructed a decision tree using the rpart package (Therneau et al. 2025): this decision tree provides conditions on the model parameters that predicts if the resulting simulation would have an AIG among the 10% highest values or not. In this algorithm, each threshold is chosen by maximizing the decrease in Gini indices between the nodes created after a split. To do that, we first label the simulations as high AIG scenario if it is in the last decile of AIG, or low AIG scenario otherwise and we re-weighted the classes with the following weights to consider class imbalances:

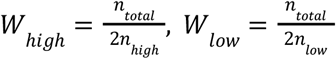

where *n*_*total*_, *n*_*high*_ and *n*_*low*_ denote respectively the number of simulations in total, with a high AIG and with a low AIG.

## Results

First, we analysed a toy example, to explore what impact asynchrony can have on simulated incidence and how we quantify it. Second, we explored the impact of intervention asynchrony in our model over a wide range of parameters, to understand under what conditions this effect matters. Finally, we analysed a series of case studies that could resemble real-life situations.

### Illustrative example

An illustrative example of the impact of intervention asynchrony on malaria incidence is presented in **Figure 2**. In this example, a lower incidence is reached when interventions are deployed synchronously in both areas. In particular, in area 1, the exact same interventions are deployed under the two scenarios considered, the only difference being the timing of the intervention in the neighboring area. Intervention asynchrony is responsible for a rebound in incidence in area 1 between year 6 and year 11, that would not have happened would area 2 had deployed the intervention at the same time. The additional incidence due to asynchrony, denoted as AIG, is represented as the area between the two curves, counted positively when the orange curve is above the black one, and negatively otherwise.

**Figure 2:**
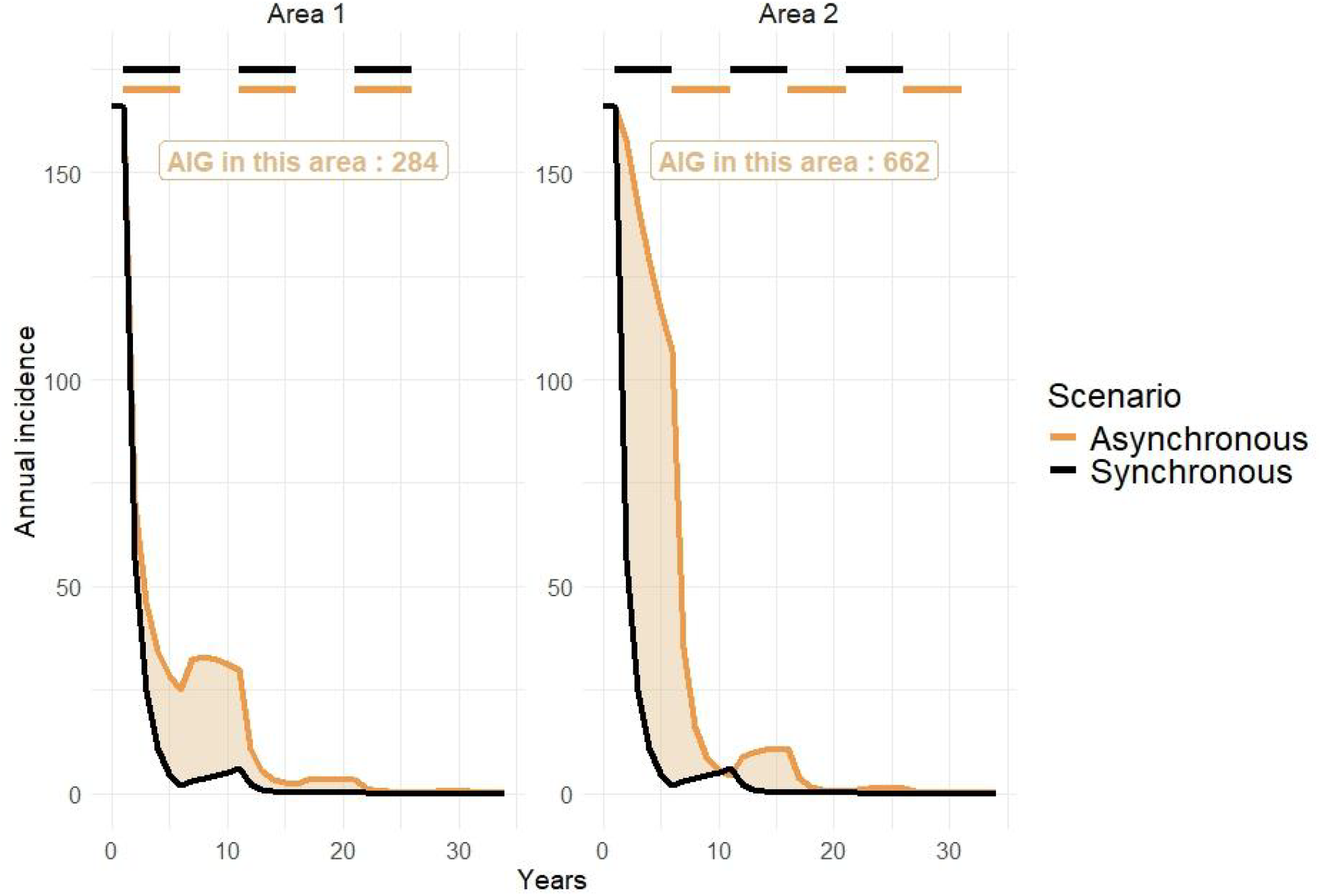
Evolution of the annual incidence in each area. The interventions start at year 1 in the simulation. The black curve represents the annual incidence in the synchronous case, while the orange curve represents the annual incidence in the asynchronous case. The orange area represents Asynchrony Induced Growth. We have an AIG by year in area 1 of 9.5, i.e an increase in annual incidence of more than 9 cases per 1000 each year over 30 years in area 1, with a total AIGR for area 1 of 62% (and an AIGR for area 2 of 147%). The first row of segments at the top represents the times when interventions are deployed in the synchronous case, while the second row represents the times when interventions are deployed in the asynchronous case. The AIG shown in the figure is the cumulated AIG in each area for the first 30 years. Parameter values used: r=1/200, t_I_ =5 years, R_0,1_ = R_0,2_ = 1. 1, ω_1_ = ω_2_ = 0. 5, p_11_ = 0. 9, p_12_ = 0. 1, p_22_ = 0. 99, p_21_ = 0. 01.

In this illustrative example, the AIG here is not considerably high, with an AIG by year of 9.4, i.e 9.4 more infections on average every year per 1000 persons in the 30 years following the beginning of the intervention due to the asynchrony, mainly concentrated in the first ten years. This example is one of a situation where both scenarios are reaching transmission interruption, but asynchrony makes it longer in time to reach this outcome (32 years later in the asynchronous case, in this case, with at least three additional 5-year intervention campaigns).

### When does intervention synchrony matter/not matter?

#### Uncertainty analysis

The distribution of AIG over all the different combination of the model parameter values is presented in **Figure 3A**. A first observation is that almost all simulations (more than 99%) have positive AIG, indicating that synchronous interventions almost always lead to a lower incidence compared to asynchronous ones. However, the distribution is concentrated in low values, with 75% of the values below 32 (meaning that in 75% of the simulations, the increase in annual incidence is below 32 infections per year per 1000 inhabitants). Therefore, in many cases, intervention asynchrony is likely to have a small epidemiological importance. Nonetheless, out of 10000 simulations, 1294 simulations correspond to an increase in annual incidence over 50 cases per 1000 people, which can be considered substantial: in these cases, asynchrony matters and has an impact. This conclusion holds also when compared to the original burden of the epidemic: out of these 1294 high burden scenarios, we have 1273 simulations where the AIGR is over 10%, i.e. after 6 cycles, the epidemic’s cumulative incidence is 10% higher in the asynchronous case compared to the synchronous case.

**Figure 3:**
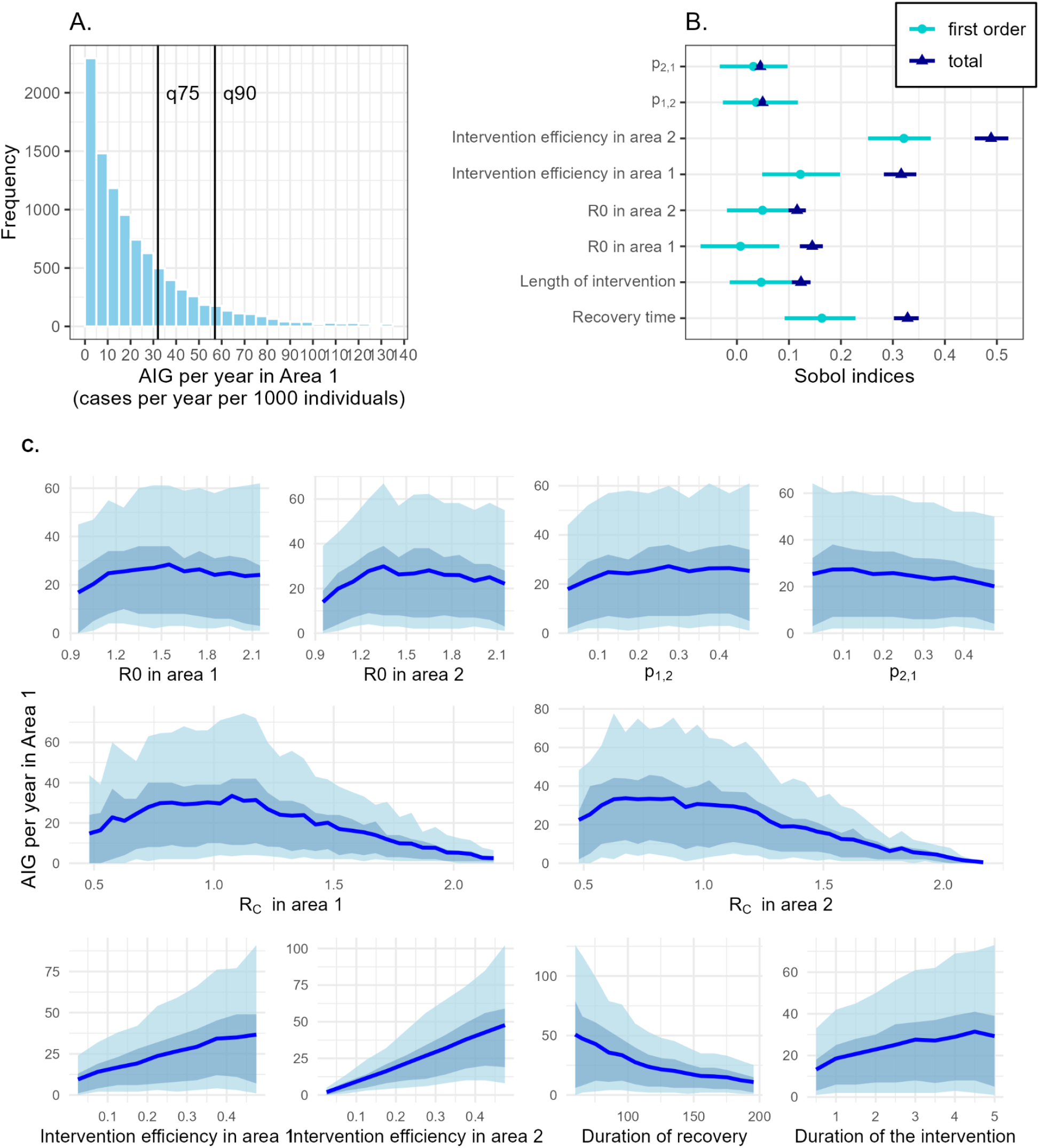
**A**. Histogram of the distribution of AIG per year in area 1 in the dataset of simulations : the first and last centile have been removed for readability (quantiles of the distribution in Appendix B). The 75 quartile and 90 quartile are represented by vertical black lines “q75” and “q90”; **B**. First-order and total Sobol sensitivity indices for model parameters.; **C**. Relationships between each parameter and AIG per year, showing median (blue line), interquartile (dark blue band) and decile ranges (light blue band).

#### Sensitivity analysis

The Sobol indices of first order and total effects are presented in **Figure 3B**. The most influential parameters are the intervention efficiencies and the recovery time for both first order effects and total effects.

Moreover, the shape of the relation between each parameter and the AIG is displayed in **Figure 3C**. The duration of recovery 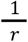 is negatively associated with the outcome, the quicker the disease is in terms of being able to reinfect a host, the more detrimental asynchrony is. AIG is an increasing function of intervention efficiency. However, the relation is different when represented in terms of *R*_0_ with control (i.e. *R*_*C*_ = *R*_0_ ×ω): we see a peak in AIG in the values around 1 (both above and below 1). We can interpret this in the following way: it is when the interventions are strong enough to reduce the epidemic strength and get closer to elimination that coordination between areas matters. The other parameters do not display substantial variability in AIG average; a slight positive correlation between the proportion of individuals moving from area 1 to area 2 (p_1_,_2_) and AIG, and a slight negative correlation between the proportion of individuals moving from area 2 to area 1 (p_2_,_1_) and AIG are however observed. This is expected, as our analysis centers on the impact of asynchrony in area 1, which serves as the counterfactual scenario. The duration of intervention impacts positively the AIG : this mainly comes from the fact that the duration of intervention is also the delay for area 2 in the beginning of interventions and also the duration of break between interventions, so for a duration of 5 years, the area 2 waits 5 years before starting its interventions, so 5 years with a full epidemic burden, and with people going to area 1 and reseeding transmission in area 1.

#### What characterizes the high impact cases?

We consider here as high the scenarios with an AIG in the last decile of the distribution of AIG, and as low the other scenarios. As shown in **Figure 4**, most of the high impact scenarios are concentrated in the low recovery time simulations, as well as the high intervention efficiency scenarios. Asynchrony has an impact when the interventions are efficient.

**Figure 4:**
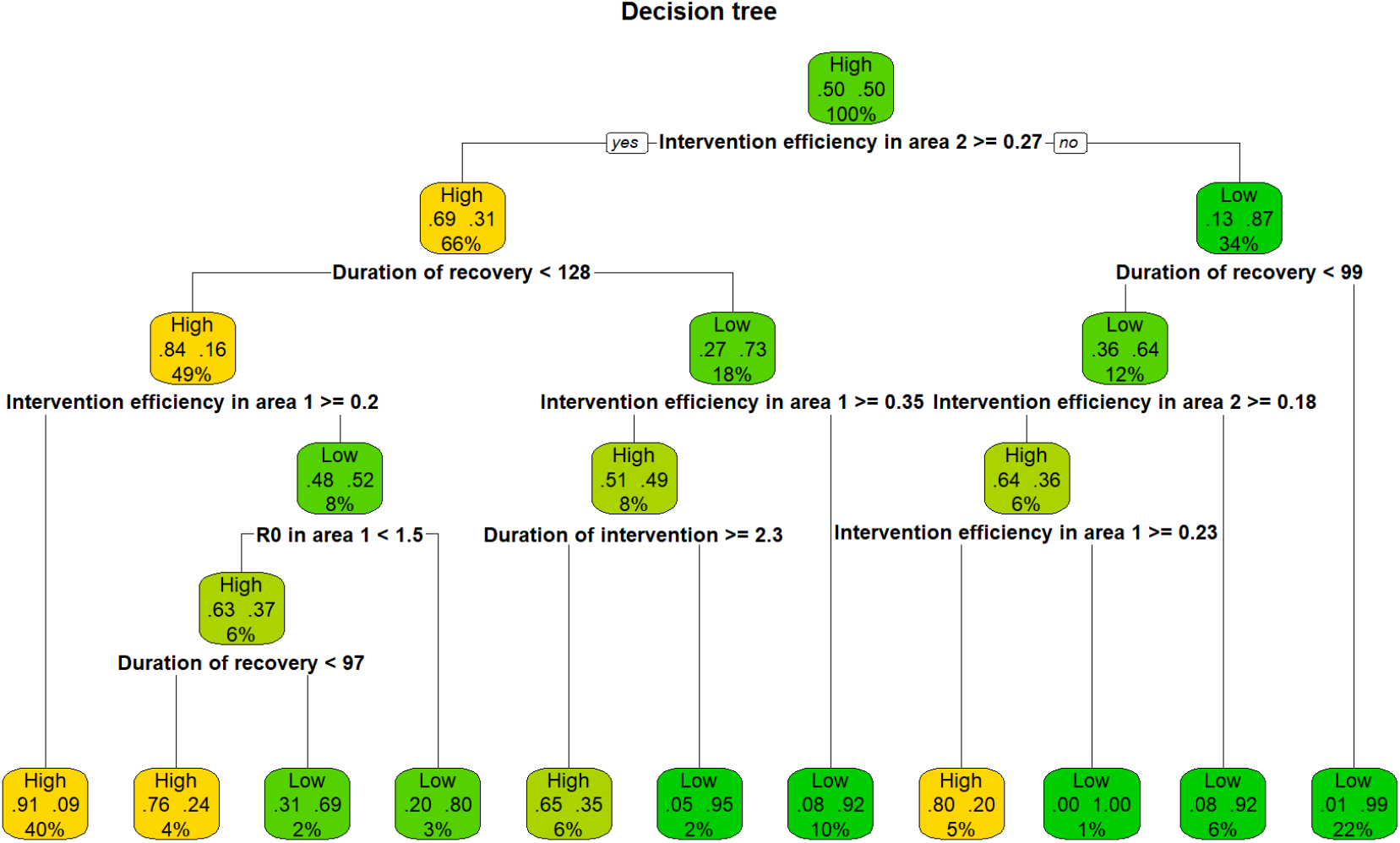
Decision tree identifying parameter thresholds predictive of high and low AIG outcomes. Each node displays the majority class (High / Low AIG) of scenarios in the node, the class proportions (the first number is the proportion of high AIG in the node, and the second one is the proportion of low AIG), and the percentage of all simulations present in the node (reweighted using weights defined in Methods section). For example, in the last row, in the first leaf, the majority class is High AIG simulations, with 91% of the simulations in this node having a label High, and 9% a label Low, and the simulations present in this node represents 40% of all the simulations.

Having built the decision tree shown in **Figure 4**, we focus on the leaves labelled high, which gives us different kinds of scenarios where the simulations have higher AIG:

- 1^st^ leaf: High-efficiency interventions in both areas with a duration of recovery lower than 128 days,
- 2^nd^ leaf: Lower-efficiency intervention in area 1 but a *R*_0,1_ lower than 1.5 (in this subspace, we have *R*_*c*, 1_ ∈(0. 72, 1, 5), so it includes scenarios with elimination or close to elimination) and a quick duration of recovery (between 60 days and 97 days),
- 3^rd^ leaf: High-efficiency interventions in both areas, but with a long duration of recovery and a long duration of intervention.
- 4^th^ leaf: Similar to the second leaf, with a recovery time below 99 days, and mid-efficiency intervention in area 2 and mid to high-efficiency intervention in area 1.

This suggests general patterns of scenarios that will be explored in the next section by looking at the actual progression of the epidemic in both cases.

## Some examples – Case studies

Figure 5 shows four scenarios selected for their plausibility with real scenarios (with extended numerical outputs presented in Appendix C).

**Figure 5:**
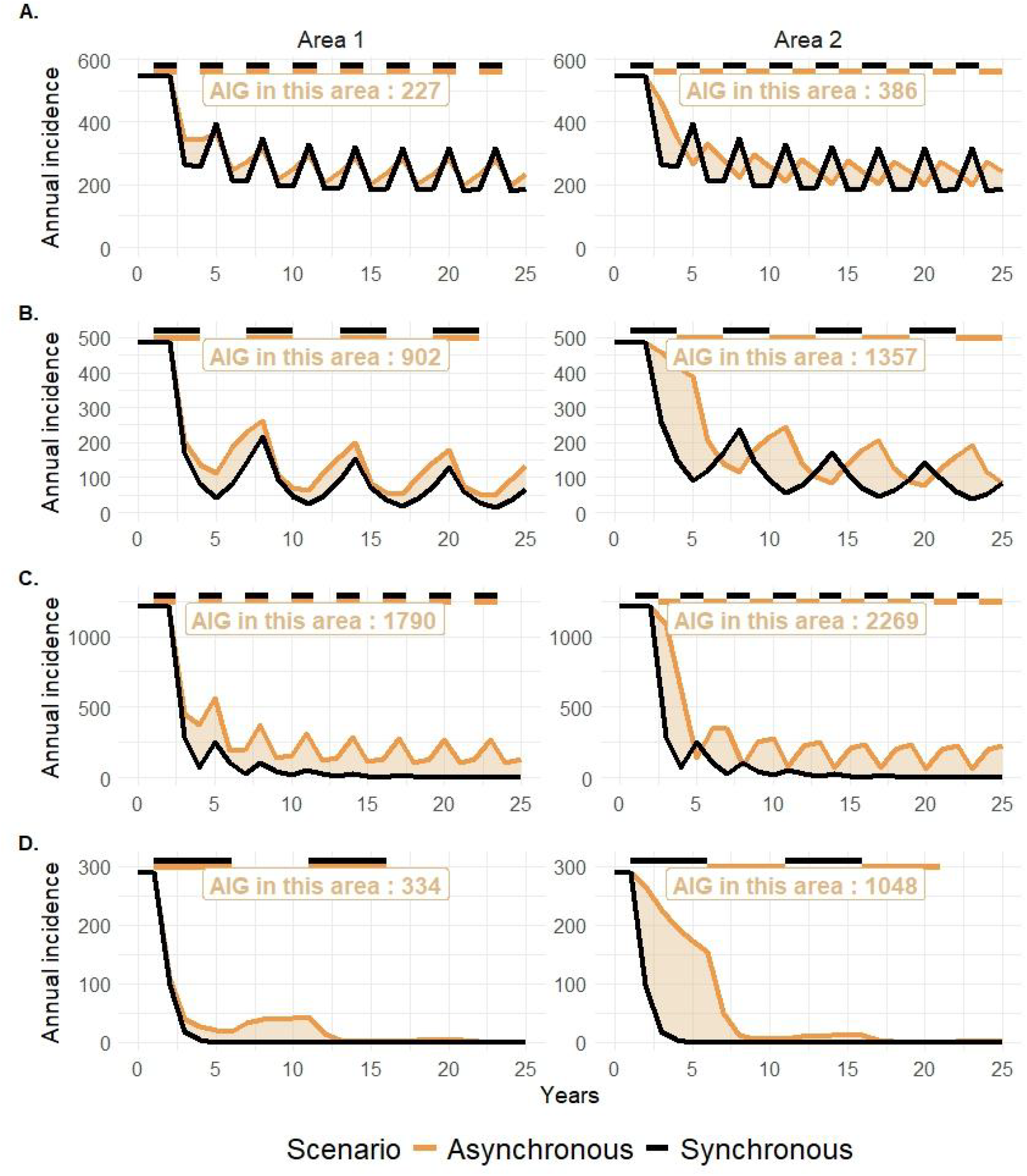
Examples of simulated scenarios reflecting plausible malaria settings. Each panel displays the time course of malaria incidence under synchronous (black) and asynchronous (orange) interventions. Parameters for each scenario : ***A***. 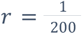, *X* _1_ = *X* _2_ = 30%, *W*_1_ = *W*_2_ = 0. 6, *t* _*I*_ = 1. 5, *p* _12_ = 0. 2, *p* _21_ = 0. 2; ***B***. 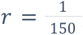, *X* _1_ = *X* _2_ = 20%, *W*_1_ = 0. 5, *W*_2_ = 0. 7, *t* _*I*_ = 3, *p* _12_ = 0. 1, *p* _21_ = 0. 01; ***C***. 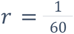, *X* _1_ = *X* _2_ = 20%, *W*_1_ = *W*_2_ = 0. 55, *t* _*I*_ = 1. 5, *p* _12_ = 0. 1, *p* _21_ = 0. 01; ***D***. 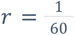, *X* _1_ = *X* _2_ = 4. 76%, *W*_1_ = *W*_2_ = 0. 7, *t* _*I*_ = 1. 5, *p* _12_ = 0. 011, *p* _21_ = 0. 03.

**Scenario A** represents two areas with high prevalence (approximately 30%), a long infectiousness duration close to the one of *Plasmodium falciparum*, and where interventions are deployed for 1.5 years and stopped for 1.5 years. Such intervention timing could correspond to insecticide-treated net (ITN) mass distribution campaigns implemented every three years, but considering that the field durability of ITNs does not exceed 1.5 years (Bertozzi-Villa et al. 2021). This scenario could represent a high burden country where ITNs distribution campaigns would be deployed independently in neighboring administrative units. Even though malaria dynamics appear to be similar in both cases, with a succession of peaks and lulls, the fact that the areas are not coordinating their interventions leads to an increase in malaria incidence. In this example, synchronizing ITN distributions in the two areas could have averted 25 more infections per 1000 inhabitants each year in area 1 in the first 9 years of the epidemic after the beginning of the first intervention, even though it represents only 0.6% of the original epidemic magnitude.

**Scenario B** represents two areas with high prevalence (approximately 30%), a long infectiousness duration close to the one of *Plasmodium falciparum*, and where interventions are deployed during 3 years and stopped for 3 years. Such interventions could correspond for example to ITN mass distribution campaigns conducted at a lower frequency due to resource constraints. In this example too, the asynchrony in intervention timing across both areas leads to a substantial increase in malaria cases. In the first area, on average, 50 cases per year per 1000 inhabitants are averted in the synchronous scenario compared to the asynchronous one, corresponding to a 35% reduction in cases over the first 18 years after the beginning of the first intervention.

**Scenario C** also represents two areas with high prevalence (approximately 20%) where interventions are deployed during 1.5 years and stopped for 1.5 years, but with a shorter infectiousness duration. This infectiousness duration could correspond to areas with high access to routine case management or to areas with high *Plasmodium vivax* endemicity such as different provinces in Papua New Guinea, where prevalence is high. Here, when deployed synchronously, interventions are efficient enough to reach interruption of malaria transmission in the long term. On the contrary, transmission interruption cannot be reached in the scenario with asynchronous interventions. The AIG by year is substantial with an increase of 199 per 1000 in annual incidence in the asynchronous scenario compared to the synchronous one in the first 9 years of the epidemic after the beginning of the first intervention.

**Scenario D** represents two areas with low prevalence (below 5%) and a short duration of infectiousness, as observed in areas with strong routine case management or with *P. vivax* transmission, such as Central America or the Greater Mekong Subregion. In this example, a strong intervention is deployed during 5 years and then interrupted during 5 years. This could represent for example an elimination campaign including Mass Drug Administration. In this example, interruption of transmission is reached in both scenarios considered, but the timelines are almost 10 years longer when interventions are asynchronised, compared to a situation where they would be deployed synchronously.

## Discussion

In this work, we explored the impact of intervention asynchrony on the success of malaria control and elimination strategies with a toy mathematical model. We found that intervention asynchrony consistently leads to an increased number of malaria cases, compared to a scenario where interventions would be deployed in a synchronized manner. Considering the long generation time characteristic of malaria dynamics, the clinical magnitude of this effect is small in most parameter sets of the studied range. However, we highlighted selected case studies resembling real-life settings, where intervention synchrony could make a significant impact on malaria dynamics, by reducing burden, enabling elimination or reducing elimination timelines. Although these cases represented only a small proportion of the explored parameter range, they could be applicable to a large number of realistic settings. This highlights the importance of exploring this topic further with data-informed examples, and of quantifying the gains in relation to the financial and operational costs required to enforce intervention synchrony.

With our simple model, we could identify the conditions under which intervention coordination matters significantly. This is particularly the case when the duration of infectiousness is shorter, resembling *P. vivax* dynamics or areas with high access to rapid effective treatment, and when the intensity of interventions is high and the intervals between them are long. Elimination programs, for example, would particularly benefit from cross-border coordination efforts. These efforts could meaningfully reduce elimination timelines, especially where resources are limited, given the costs of extending elimination timelines and the uncertain nature of funding. Initiatives such as MOSASWA (Moonasar et al., 2016), which implements elimination interventions across Southern Mozambique, South Africa and Eswatini are very valuable, and should therefore not only emphasize the location of interventions across different countries but also their relative timing.

In our model, the duration of infectiousness was highlighted as a crucial factor for intervention asynchrony to lead to high case surges. Untreated malaria can be transmitted over very long periods of time and this contributed to the smaller impact of intervention asynchrony in many tested scenarios. However, this means intervention asynchrony may have a higher importance for diseases with shorter generation time, such as respiratory viruses. This was already shown for lockdowns (Ruktanonchai et al. 2020) implemented as a response to Covid-19 and could also have importance for other diseases.

We also restricted our analysis to the deployment of new interventions in endemic settings. Other scenarios could also be explored, such as the impact of intervention asynchrony in non-equilibrium epidemic settings, the impact of asynchrony in intervention removal in areas where control measures are already in place or the impact of asynchrony as a random event, to represent budget cuts or change of governments or regimes.

Our conceptual framework, as well as the provided computer code, could be readily expanded to study such situations. Our metric, the Asynchrony Induced Growth (AIG), enabled us to explore this phenomenon across a wide range of parameter combinations in a standardized fashion. Coupled with sensitivity analyses and machine learning tools, we could identify the conditions under which asynchrony can make a difference of public health relevance. In future work, this approach can be used with more complex models or scenarios to explore more broadly the concept of intervention asynchrony on disease dynamics. It could also be used to identify ways to do damage limitation in situations where interventions will realistically have to occur asynchronously, i.e comparing two plausible asynchronous cases to find out which one would be most appropriate for the situation.

Nevertheless, this work has some limitations. It relies on an SIS model with two patches and some key characteristics of malaria transmission were not included, such as explicit vector dynamics, immunity in hosts or the relapse mechanism for *P. vivax*. Therefore, further work could focus on expanding this proof of concept into real-life settings, by formally calibrating more realistic models to available data. Moreover, in our statistical analysis, we considered that all scenarios are equiprobable, when the set of realistic scenarios might be much smaller. Another limitation is the absence of consideration of seasonality in the model. Seasonal variations are a key feature of malaria transmission dynamics in most environments and such fluctuations were highlighted by (Kortessis et al. 2025) as an important driver of spatio-temporal heterogeneity in ecological populations. Climate drivers may be either uniform or diverse in cross-border areas. Therefore, depending on the local context, seasonality may increase or reduce the spatio-temporal heterogeneity due to intervention asynchrony. Studying such considerations using real data would also be beneficial.

In conclusion, this study highlights the role of intervention asynchrony in shaping malaria transmission dynamics. While our results indicate that asynchronous deployment of interventions generally increases malaria burden compared to synchronized implementation, the overall magnitude of this effect remains modest in most endemic settings due to the long infectious period of malaria. Nonetheless, we identified specific scenarios -particularly those resembling *P. vivax* transmission, high intervention intensity, or long intervention intervals - where synchrony could significantly accelerate elimination and reduce case numbers. These findings underscore the importance of considering temporal coordination, especially in cross-border elimination campaigns or settings with short infectious durations. Ultimately, our work provides a conceptual and computational foundation for further exploration of intervention timing in more realistic, data-driven models, with broader implications for optimizing public health strategies against malaria and other infectious diseases.

## Data Availability

All code necessary to reproduce the figures and conclusions of the present study are provided at : https://github.com/SwissTPH/asynchrony_simulations.

## Acknowledgements

Computational analyses were performed using resources provided by the SciCORE scientific computing center at the University of Basel.

This work was supported in whole or in part by the Gates Foundation [Grants INV-072375 and INV-068864]. The conclusions and opinions expressed in this work are those of the author(s) alone and shall not be attributed to the Foundation. Under the grant conditions of the Foundation, a Creative Commons Attribution 4.0 License has already been assigned to the Author Accepted Manuscript version that might arise from this submission.

A special thanks to Nicholas Putney for his help with the use of the odin package.

## Data and code availability

All code necessary to reproduce the article’s figures and conclusions are provided at : https://github.com/SwissTPH/asynchrony_simulations.

## Appendix A

We write 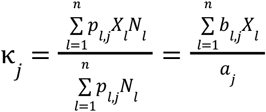 with *b*_*l,j*_ = *p*_*l,j*_ *N*_*j*_ and 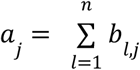. We denote by *F*_*i*_ the right-hand side of (1) :

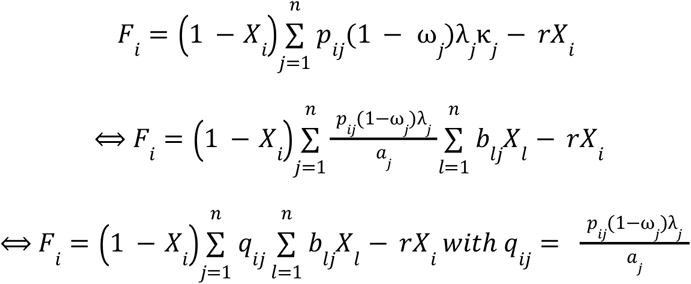

Let’s F be the following vectorial function:

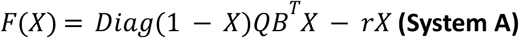

where:

- 1 − *X* = (1 − *X*_1_, 1 – *X*_2_, …, 1 − *X*_*n*_ )∈ *R*^*n*^,
- *Diag*(*Y*) for a vector *Y* = (*Y*_1_, …, *Y*_*n*_ )∈ *R*^*n*^ is the diagonal matrix *D*∈*M*_*n*_ (*R*) with *D*_*ii*_ = *Y*_*i*_,
- *Q* = (*q*_*ij*_)_1≤*i,j*≤*n*_ ∈ *M*_*n*_ (*R*) and
- *B* = (*b*_*ij*_)_1≤*i,j*≤*n*_ ∈ *M*_*n*_ (*R*).

The Jacobian of *F* is given by: *J*(*X*) = *Diag*(1 − *X*)*QB*^*T*^ − *rI*_*n*_ − *Diag*(*QB*^*T*^ *X*).

We then have the following differential equation 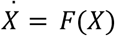. This equation is defined on the unit cube *K* = [0, 1]^*n*^ . We’ll use Corollary 3.2 of (Zhao and Jing 1996) to conclude that system (1) admits a unique positive equilibrium *X*^*\**^ (i.e. an endemic equilibrium) that is globally asymptotically stable with respect to 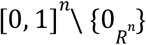.

The conditions to fulfill are :

1. *F* is cooperative on *K* and the jacobian of *F* is an irreducible matrix for every *X* in the interior of *K*.
2. *F*(0)=0 and K is positively invariant.
3. *F* is strictly sublinear on *Int*(*K*).
4. *J*(0) is unstable, hence its spectral abscissa (written *s*(*J*(0))) is strictly positive.

We check every condition, with condition 4 checked only for *n* = 2 as it is the condition in which we place our study :

1. Since 1 − *X*≥0, *J*(*X*) is a Metzler matrix, i.e. a matrix with non-negative non-diagonal coefficients (*J*(*X*)_*ij*_≥0 for all *i*≠*j*). Therefore, **F is cooperative on** *K* and it can also be shown that *J*(*X*) **is an irreducible matrix for every** *X*∈*Int*(*K*).
2. We have trivially that *F*(0) = 0. If *X*_*i*_ = 0,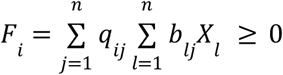 and if *X*_*i*_ = 1, then *F*_*i*_ =− *rX*_*i*_ ≤ 0. Hence, *K* **is positively invariant** which means that if the initial condition belongs to K then the solution starting from this initial condition will stay in *K* for all *t*≥0. This implies that **all the solutions are bounded**.
3. For *n* = 2, we have: *J*(*x*_1_, *x*_2_ ) =[(1 − 2*x*_1_ )(*q*_11_ *b*_11_+ *q*_12_ *b*_12_ ) − *c*_1_ *x*_2_ − *r c*_1_ (1 − *x*_1_ ) *c*_2_ (1 – *x*_2_ ) (1 − 2*x*_2_ )(*q*_21_ *b*_21_+ *q*_22_ *b*_22_ ) – *c*_2_ *x*_1_ − *r* ] with *c*_1_ = *q*_11_ *b*_21_ + *q*_12_ *b*_22_ and *c*_2_ = *q*_21_ *b*_11_ + *q*_22_ *b*_12_ . For α∈(0, 1) and *X*∈*Int*(*K*), we have *F*(α*X*) = *Diag*(1 − α*X*)*QB*^*T*^ α*X* − *r*α*X* > α*F*(*X*) and so, *F* **is strictly sublinear on** *Int*(*K*).
4. Moreover, *J*(0) = *QB*^*T*^ − *rI*_*n*_ = [(*q*_11_ *b*_11_ + *q*_12_ *b*_12_ ) − *r c*_1_ *c*_2_ (*q*_21_ *b*_21_ + *q*_22_ *b*_22_ ) − *r* ]. We define *R*_0_ as *s*(*QB*^*T*^ /*r*). We have *s*(*J*(0)) = *r*(*R*_0_ − 1). Then, when *R*_0_ > 1, **this matrix is unstable**.

Hence, we have all the conditions to use Corollary 3.2 of (Zhao and Jing 1996) and get the wanted result.

## Appendix B

Because we have a very concentrated distribution, we chose to indicate the 1st and last centile additionally to the quartiles.

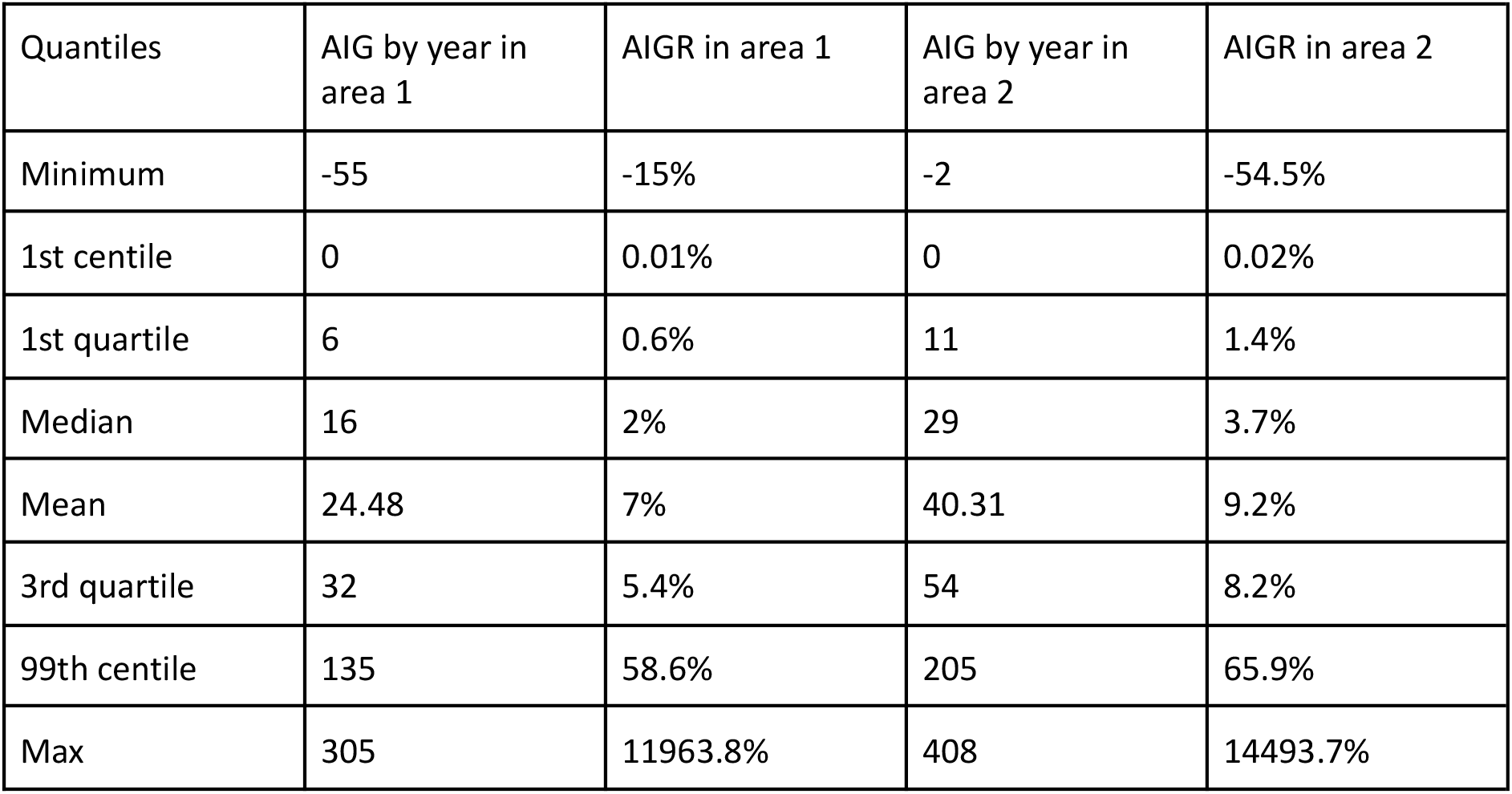

## Appendix C

| Scenario | Duration of an intervention | Number of years* | AIG by year in area 1 | AIGR in area 1 | AIG by year in area 2 | AIGR in area 2 |
| --- | --- | --- | --- | --- | --- | --- |
| Scenario A | 18 months | 9 years | 25 | 0.6% | 43 | 1% |
| Scenario B | 3 years | 18 years | 50 | 35.6% | 75 | 38.4% |
| Scenario C | 18 months | 9 years | 199 | 52.8% | 252 | 49.9% |
| Scenario D | 5 years | 30 years | 11 | 47.6% | 35 | 105.8% |

\*(for computation of AIG by year, based on the duration multiplied by the number of cycles, i.e 6)

